# Characteristics, service use and mortality of clusters of multimorbid patients in England: a population-based study

**DOI:** 10.1101/19000422

**Authors:** Yajing Zhu, Duncan Edwards, Jonathan Mant, Rupert A Payne, Steven Kiddle

## Abstract

**Background:** Multimorbidity is one of the principal challenges facing health systems worldwide. To help understand the changes to services and policies that are required to deliver better care, we used a novel approach to investigate which diseases co-occur and how combinations are associated with mortality and service use.

**Methods:** Linked primary and secondary care electronic health records contributed by 382 general practices in England to the Clinical Practice Research Datalink (CPRD) were used. The study included a representative set of multimorbid adults (18 years old or more) with two or more long-term conditions (N=113,211). A random set of 80% of the multimorbid patients (N=90,571) were stratified by age and clustered using latent class analysis. Consistency between obtained disease profiles, classification quality and associations with demographic characteristics and primary outcomes (hospitalisation, polypharmacy and mortality) was validated in the remaining 20% of multimorbid patients (N=22,640).

**Findings:** We identified twenty patient clusters across four age strata. The clusters with the highest mortality comprised psychoactive substance and alcohol misuse (aged 18-64), coronary heart disease, depression and pain (aged 65-84) and coronary heart disease, heart failure and atrial fibrillation (aged 85+). The clusters with the highest service use coincided with those with highest mortality for people aged over 65. For people aged 18-64, the cluster with the highest service use comprised depression, anxiety and pain. The majority of 85+ year olds multimorbid patients belonged to the cluster with the lowest service use and mortality for that age range. Pain featured in thirteen clusters.

**Interpretation:** This work has highlighted patterns of multimorbidity that have implications for health services. These include the importance of psychoactive substance and alcohol misuse in people under the age of 65; of co-morbid depression and coronary heart disease in people aged 65-84, and of cardiovascular disease in people aged 85+.

**Funding:** UK Medical Research Council

**Research in context:** *Evidence before this study:* We searched PubMed using the keyword list “multimorbidity, co-morbidity, disease patterns, clusters, service use, long-term conditions, chronic conditions” for studies published in English. We also reviewed multiple systematic reviews of multimorbidity patterns, and the policy report of multimorbidity research issued by the Academy of Medical Sciences in 2018. Most studies have focused on older populations (aged 60+) and often used a small list of long-term conditions. Multimorbidity clusters composed of more than two conditions have not been well profiled mostly due to non-representative and small samples. There is substantial heterogeneity in the number of conditions considered (often less than 20) and in the statistical methods. Most studies focused on grouping diseases rather than patients, making it less straightforward to relate patients to outcomes in order to facilitate patient-centred care.

*Added value of this study:* This study is the first to describe and validate clusters of multimorbid patients across the adult lifecourse using a patient-centred probabilistic clustering approach. This leads to a more nuanced understanding of the relationship between age, multimorbidity and mortality and new insights into the importance of different clusters. For example, contrary to perceived wisdom, we show that the majority of 85+ year old multimorbid patients belong to a cluster with relatively low service use and mortality for that age group. We identify a cluster of younger multimorbid patients with psychoactive substance misuse that have a mortality rate 18 times higher than their non-multimorbid peers.

*Implications of all the available evidence:* We have validated and added to the list of disease combinations where tailored approaches could be attempted to better manage multimorbid patients and develop effective interventions. For example, the high mortality of younger multimorbid patients with psychoactive substance misuse might be reduced by addressing risk factors (e.g. drug use, smoking, deprivation) which are amenable to intervention.

## Introduction

As a result of improved life expectancy and ageing populations, a growing number of individuals are living with multimorbidity, i.e. more than one long-term condition^1,2^. Multimorbidity has been recognised as a global challenge for health care management^3^ and it is estimated by the Health Foundation that 14 million individuals in England have multimorbidity, with over a third of these having more than four long-term conditions^4^. Patients with multimorbidity also account for the majority of primary care consultations and hospitalisations in the UK^5^. However, current clinical specialities, guidelines, quality improvement strategies and quality of care metrics are organised around single diseases^1^ and treatments of multiple conditions are rarely coordinated, resulting in insufficient or even conflicting care^6^.

Patients with multimorbidity have a diverse range of diseases, needs and outcomes^4,5,7^. Identifying and characterising groups of multimorbid patients that share similar patterns of long-term conditions might facilitate an improvement in their healthcare. For example, such an approach might aid the development of effective strategies for early diagnosis and prevention of multimorbidity, and allow for a better design and delivery of targeted interventions^1,8^. Several systematic reviews have found common multimorbidity clusters involving cardiovascular-metabolic conditions, mental health and musculoskeletal disorders^9,10^.However, existing evidence has important limitations. Most previous studies have focused on older populations (aged 60+); few have provided age-stratified clusters^9–11^, leaving scarce evidence for the younger multimorbid population. Second, multimorbidity clusters composed of more than two conditions have not been well profiled mostly due to non-representative and smaller samples^1,9,10^. Third, there is substantial heterogeneity in the number of conditions considered (often less than 20) and in the statistical methods. Most studies focused on grouping diseases rather than patients, where each disease can only go into one cluster and so it is not straightforward to relate patients to outcomes in order to facilitate patient-centred policy-making^12^. Commonly used clustering methods were exploratory approaches such as factor analysis and hierarchical clustering^9,13^, where results were highly sensitive to the subjective choice of metrics^14^. Finally, the validity and generalisability of cluster solutions in new samples is important for decision-making but is often ignored in the current literature^9,10^.

This study aims to identify, validate and study the outcomes of age-stratified clusters of multimorbid adult patients in a large representative sample of UK patients. Towards this end we used a comprehensive list of 38 long-term conditions^5^ and a robust model-based probabilistic approach, latent class analysis^14^.

## Methods

### Data source

Our analysis used the Clinical Practice Research Datalink (CPRD)-GOLD database where anonymised and longitudinal primary care clinical data are contributed by UK general (family) practices (GP) who use the Vision health record system^15^. CPRD has been validated to be representative of the UK population for age, sex, and ethnicity^15,16^. Patients’ GP records were linked with hospitalisation data (Hospital Episodes Statistics, HES), all-cause mortality data (Office for National Statistics, ONS) and area socioeconomic deprivation data (Index of Multiple Deprivation, IMD); these linked data were available for approximately 75% of CPRD English practices. The protocol for this study (16_057RA2) was approved scientifically and ethically by the CPRD Independent Scientific Advisory Committee.

### Study population

Data on a random selection of individuals were acquired from CPRD (the same individuals studied in Cassell et al.,^5^). Patients aged 18 years and above with valid registered-status in a practice with data classified by CPRD as “up-to-standard” in January 2012^15^ were included in the study. We chose the year 2012 to allow complete ascertainment of five-year mortality. Additionally, we required that their practice allowed linkage to ONS, IMD and HES, resulting in the inclusion of only English practices.

### Patient and public involvement

There was no patient or public involvement in this study.

### Statistical software

Data analysis was performed in R 3.4.4. R package names are given in the following sections where appropriate (*in brackets and italics*), for example memory efficient packages were used to extract data for analysis in R (*ff, CALIBERdatamanage*). For transparency and reproducibility, all analysis scripts and code lists are available from https://github.com/Kiddle-group.

#### Definition of patient characteristics, morbidities and outcomes

Morbidities in this study were defined as binary variables (present or not) based on the classification of LTCs in primary care developed by Barnett et al.,^7^ This taxonomy attempted to include all conditions “likely to be chronic (defined as having significant impact over at least the most recent year) and with significant impact on patients in terms of need for chronic treatment, reduced function, reduced quality of life, and risk of future morbidity and mortality”, and was developed for use in UK primary care electronic health record research and has been adapted for use in CPRD^5^. The specific definitions for each LTC is based on the UK Read code system and electronic prescription data coded using CPRD’s prodcode, giving a total of 38 LTCs. (https://www.phpc.cam.ac.uk/pcu/cprd_cam/codelists/v11/). The LTCs used in this study largely match the only other large sample size UK multimorbidity cluster study^17^.

Two sets of outcome variables related to service use and mortality were defined. NHS service utilisation or treatment burden was measured by three variables over the 12-month period after January 2012: primary care consultations (consultations with any clinician in the primary care team), the number of all-type hospitalisation spells (defined by discharge dates) and the amount of repeat prescriptions (at least four times in a year by counting the unique British National Formulary (BNF) codes). All-cause mortality at two and five years was extracted from ONS data.

Patient characteristics that were considered in this study include gender, age groups (stratified into 18-44, 45-64, 65-84 and 85+ years) in 2012, last recorded pre-2012 body mass index (BMI), last recorded pre-2012 smoking status (current, never and ex-smokers) and socioeconomic status measured by IMD in quintiles (1 for the least deprived and 5 for the most).

### Statistical analysis

This study aims to identify clusters of multimorbid patients using patterns of co-existing long-term conditions. We used latent class analysis (LCA) (*poLCA*) to assign all patients to non-overlapping clusters (i.e. each patient is assigned to only one cluster) in a data-driven fashion^18,19^. Compared to other exploratory clustering methods (e.g. factor analysis, hierarchical clustering^10,20^), LCA is a model-based probabilistic clustering approach that is not sensitive to rotation of factors and does not require any subjective choice of “distance measures” for multimorbidity patterns^14,21^.This greatly enhances the reproducibility and stability of the latent class solutions. Clustering patients rather than diseases allows diseases to belong to multiple clusters and more naturally allows the characteristics and outcomes of clusters to be studied. As a result, each derived patient cluster has a unique and probabilistic multimorbidity phenotype profile where members do not necessarily need to have all conditions.

Guided by simulation studies^22^, the optimal number of latent classes was decided using a combination of statistics (Bayesian Information Criteria (BIC), sample-sized adjusted BIC, log-likelihood ratio test, entropy for classification quality) and clinical judgment. Within our datasets conditions are present (i.e. recorded) or not by definition, and so missing data methods were not needed for cluster analysis. More details on a technical review of commonly used clustering methods, the LCA methodology and application of selection statistics are provided in supplementary material section 3.

To account for the different nature of multimorbidity clusters at different ages, four age strata (18-44, 45-64, 65-84, 85+ years) were chosen. We derived the cluster solution and performed post-hoc statistical tests in a stratified (by age strata) random sample of the multimorbid population that contained 80% of the patients (i.e. training set). Separate LCAs were performed for each stratum, and each patient allocated to a single multimorbidity cluster. For ease of interpretation, clusters were labelled by their three most distinctive conditions whose difference in prevalence between cluster and age strata were the highest (see supplementary table 1 for full details of conditions). To quantify the association between outcomes, multimorbidity clusters and patient demographics, generalised linear models were fitted (see supplementary material section 5). Individuals with missing data for last pre-2012 recording of smoking status and BMI represented a small percentage (<5%) of the population, and so were excluded from the generalised linear models.

**Table 1.** Demographic characteristics of the whole population (N=391,669). For ordinal variables median, first (Q1) and third (Q3) quartiles are reported. For categorical variables, counts and percentages are reported.

| Demographics | N (%) | # Morbidities<br>Median<br>[Q1 - Q3] | Multimorbid patients<br>(%) |
| --- | --- | --- | --- |
| All patients | 391,669 (100) | 1 [0-2] | 28.9 |
| Gender |  |  |  |
| Male | 192,929 (49.3) | 0 [0-2] | 26.0 |
| Female | 198,740 (50.7) | 1 [0-2] | 31.7 |
| Age group (years) |  |  |  |
| 18-24 | 32,007 (8.2) | 0 [0-0] | 4.7 |
| 25-34 | 60,501 (15.4) | 0 [0-1] | 7.9 |
| 35-44 | 68,688 (17.5) | 0 [0-1] | 13.1 |
| 45-54 | 74,734 (19.1) | 0 [0-1] | 20.5 |
| 55-64 | 60,323 (15.4) | 1 [0-2] | 34.4 |
| 65-74 | 49,427 (12.6) | 2 [1-3] | 53.6 |
| 75-84 | 31,262 (8.0) | 3 [1-4] | 73.6 |
| 85+ | 14,727 (3.8) | 4 [2-5] | 83.6 |
| Socioeconomic status |  |  |  |
| 1 (least deprivation) | 90,730 (23.2) | 1 [0-2] | 27.1 |
| 2 | 87,734 (22.4) | 1 [0-2] | 28.5 |
| 3 | 81,569 (20.8) | 1 [0-2] | 29.1 |
| 4 | 71,424 (18.2) | 1 [0-2] | 29.3 |
| 5 (greatest deprivation) | 60,212 (15.4) | 1 [0-2] | 31.4 |
| Independence test for demographics and multimorbidity |  |  |  |
| Gender vs Multimorbidity: $\chi^2$ (1)= 1573.4, $p<0.001$ | | | |
| Age group vs Multimorbidity: $\chi^2$ (7) =100340.1, $p<0.001$ | | | |
| IMD vs Multimorbidity: $\chi^2$ (4) =337.4, $p<0.001$ | | | |

#### Assessment of the stability of morbidity clusters

To assess the stability of age-stratified multimorbidity clusters, LCAs were repeated in the remaining 20% of the population (i.e. test set), fixing the number of clusters to match that learned from the training set^23^. We employed three methods to indirectly validate our cluster solutions (a direct approach was not possible as clusters were unobserved). First, to check the consistency between disease profiles for 38 LTCs in the training and test sets, each cluster in the test set was matched (using two criteria for robustness) with a corresponding cluster in the training set. Matched cluster pairs were selected such that Jensen–Shannon distance^24^ (JSD, a measure of the divergence between disease profiles) is the smallest and the bivariate Pearson’s correlation coefficient^25^ (the degree to which two disease profiles covary) is the highest (supplementary tables 4a, 4b). Second, entropy measures^22^ (for classification quality) computed in the training and test sets were expected to be similar.

Finally, stability was further assessed by observing in the training and test sets similar associations (in terms of size, direction and statistical significance) between clusters, patient demographics and outcome variables. For more details, see supplementary material section 4.

#### Role of the founding source

The funder is not involved in study design, data collection, analysis, interpretation, report writing and submission. YZ and SJK had full access to all the data in the study and had final responsibility for paper submission.

## Results

### Characteristics of the study population

A total of 391,669 patients were included in the study, of which 49% and 22% had none or only one long-term condition respectively (see Table 1 for patient demographics). Females, older individuals and those from areas of greater socioeconomic deprivation had a higher prevalence of multimorbidity.

Among the multimorbid patients (N=113,211, 29%), all unique combinations of conditions were less than 1% prevalent in the total population with the most prevalent 20 containing only pairs of conditions (supplementary table 2). This together with the large number of unique combinations of conditions (supplementary table 3) indicated that multimorbidity patterns were highly heterogeneous. Stratified by age strata, Table 2 shows that multimorbidity in the younger population (18-44) was more common in areas with greater deprivation while for older groups, the pattern is reversed.

**Table 2.** Characteristics of multimorbid patients (N=113,211). For continuous and ordinal variables, median, first (Q1) and third (Q3) quartiles are reported. For categorical variables, counts and percentages are reported.

| Demographic characteristics |  | Across age strata<br>(N = 113,211) | Age 18-44 years<br>(N = 15,306) | Age 45-64 years<br>(N = 36,097) | Age 65-84 years<br>(N = 49,494) | Age 85+ years<br>(N = 12,314) |
| --- | --- | --- | --- | --- | --- | --- |
| Female (%) |  | 63,072 (56) | 9,422 (62) | 19,690 (55) | 25,952 (52) | 8,008 (65) |
| # morbidities: Median [Q1 - Q3] |  | 3 [2-4] | 2 [2-3] | 2 [2-3] | 3 [2-4] | 4 [3-6] |
| Age: Median [Q1 - Q3] |  | 66 [53-77] | 37 [30-41] | 56 [51-61] | 74 [69-79] | 89 [86-91] |
| BMI |  |  |  |  |  |  |
|  | Median [Q1 - Q3] | 27 [24-31] | 26 [23-31] | 28 [25-33] | 27 [24-31] | 25 [22-28] |
|  | Missing (%) | 5,788 (5) | 1,571 (10) | 1,462 (4) | 1,540 (3) | 1,215 (10) |
| Smoking status (%) |  |  |  |  |  |  |
|  | Current smoker | 21,586 (19) | 5,641 (37) | 9,140 (25) | 6,160 (12) | 645 (5) |
|  | Never smoker | 55,145 (49) | 6,789 (45) | 17,040 (47) | 24,042 (49) | 7,274 (59) |
|  | Ex smoker | 36,265 (32) | 2,821 (18) | 9,869 (27) | 19,251 (39) | 4,324 (35) |
|  | Missing | 215 (0.19) | 55 (0.36) | 48 (0.13) | 41 (0.08) | 71 (0.58) |
| Index of multiple deprivation in quintiles (%) |  |  |  |  |  |  |
|  | 1 (least deprivation) | 24,624 (22) | 2,647 (17) | 7,333 (20) | 11,722 (24) | 2,922 (24) |
|  | 2 | 25,027 (22) | 2,738 (18) | 7,668 (21) | 11,667 (24) | 2,954 (24) |
|  | 3 | 23,700 (21) | 2,969 (19) | 7,352 (20) | 10,612 (21) | 2,767 (22) |
|  | 4 | 20,934 (18) | 3,316 (22) | 6,862 (19) | 8,645 (17) | 2,111 (17) |
|  | 5 (greatest deprivation) | 18,926 (17) | 3,636 (24) | 6,882 (19) | 6,848 (14) | 1,560 (13) |

**Table 3.** Descriptions of the derived clusters of multimorbid patients for each age strata. Clusters are ordered by sizes from the largest to the smallest. Key conditions are the three estimated to be most distinctive in the cluster (where the difference between within-cluster prevalence and prevalence in age strata is the largest). For the number of morbidities, median, first (Q1) and third (Q3) quartiles are reported. For other categorical variables, percentages are reported. Greater deprivation denotes top 40% of IMD (categories 4 and 5).

| Three key conditions (prevalence) |  | Patients | # morbidities | Female | Greater deprivation | Current smokers |
| --- | --- | --- | --- | --- | --- | --- |
| Lead condition (%) | Subsidiary conditions (%) | (%) | (Median [Q1-Q3]) | (%) | (%) | (%) |
| **Age 18-44 years** |  |  |  |  |  |  |
| Depression (100%) | Anxiety (41%), Pain (31%) | 32 | 2 [2-3] | 66 | 50 | 46 |
| Pain (36%) | Hearing loss (30%), Hypertension (23%) | 23 | 2 [2-3] | 52 | 46 | 27 |
| Asthma (100%) | IBS (26%), Depression (20%) | 20 | 2 [2-3] | 63 | 41 | 29 |
| IBS (100%) | Depression (29%), Hearing loss (21%) | 18 | 2 [2-3] | 77 | 37 | 28 |
| PSM (75%) | Alcohol (42%), Depression (24%) | 7 | 2 [2-3] | 28 | 63 | 76 |
| **Age 45-64 years** |  |  |  |  |  |  |
| Hypertension (76%) | Diabetes (37%), Pain (25%) | 37 | 2 [2-3] | 42 | 38 | 20 |
| IBS (40%) | Hearing loss (29%), Pain (28%) | 24 | 2 [2-3] | 64 | 29 | 20 |
| Depression (93%) | Pain (53%), Anxiety (31%) | 22 | 3 [2-5] | 68 | 46 | 35 |
| Asthma (100%) | Pain (24%), COPD (16%) | 12 | 2 [2-3] | 61 | 35 | 20 |
| Alcohol (62%) | PSM (42%), Pain (28%) | 4 | 3 [2-4] | 31 | 57 | 63 |
| **Age 65-84 years** |  |  |  |  |  |  |
| Hypertension (100%) | Diabetes (31%), Pain (27%) | 41 | 3 [2-4] | 54 | 30 | 10 |
| Hearing loss (40%) | Prostate disorder (21%), IBS (3%) | 22 | 3 [2-4] | 48 | 25 | 9 |
| Depression (56%) | Pain (56%), Anxiety (23%) | 14 | 4 [3-5] | 72 | 33 | 15 |
| CHD (54%) | Diabetes (32%), Atrial fibrillation (29%) | 11 | 4 [3-5] | 30 | 33 | 12 |
| COPD (57%) | Asthma (49%), Pain (33%) | 8 | 3 [2-5] | 50 | 40 | 24 |
| Pain (81%) | CHD (53%), Depression (45%) | 5 | 7 [7-9] | 54 | 43 | 16 |
| **Age 85+ years** |  |  |  |  |  |  |
| Hypertension (72%) | Hearing loss (39%), Diabetes (18%) | 58 | 3 [2-4] | 61 | 30 | 5 |
| Pain (64%) | Depression (41%), Constipation (24%) | 23 | 5 [4-6] | 80 | 30 | 5 |
| CHD (61%) | Atrial fibrillation (53%), Heart failure (49%) | 11 | 7 [6-8] | 60 | 30 | 4 |
| Asthma (48%) | COPD (48%), Pain (44%) | 8 | 5 [4-6] | 59 | 30 | 8 |

Notes: IBS=Irritable bowel syndrome, PSM = Psychoactive substance misuse (not alcohol), CHD = Coronary heart disease, COPD = Chronic Obstructive Pulmonary Disease

**Table 4.** Mortality and health service utilisation by patient clusters in each age strata. Clusters are ordered by the highest to lowest mortality. The non-multimorbid cluster contains patients with zero or only one long-term condition. The number of GP consultations, hospitalisations and repeat prescriptions (by counting the number of unique BNF codes that were in repeated prescriptions at least four times), are measured in one year after January 2012. Both mean and median are reported because they highlight different aspects of skewed distributions, especially in relation to hospitalisations and prescriptions.

| Three key conditions (prevalence) |  | 2-year mortality (%) | 5-year mortality (%) | # GP contacts in a year (Mean, Median [Q1-Q3]) | # hospitalisations in a year (Mean, Median [Q1-Q3]) | # unique medicine on repeat prescriptions in a year (Mean, Median [Q1-Q3]) |
| --- | --- | --- | --- | --- | --- | --- |
| Lead condition (%) | Subsidiary conditions (%) |  |  |  |  |  |
| **Age 18-44 years** |  |  |  |  |  |  |
| PSM (75%) | Alcohol (42%), Depression (24%) | 1.8 | 3.9 | 10.7, 7 [1-16] | 0.4, 0 [0-0] | 1.3, 0 [0-2] |
| Pain (36%) | Hearing loss (30%), Hypertension (23%) | 1.0 | 2.7 | 11.9, 9 [3-17] | 0.6, 0 [0-0] | 2.5, 1 [0-4] |
| Depression (100%) | Anxiety (41%), Pain (31%) | 0.9 | 1.8 | 14.5, 12 [5-20] | 0.4, 0 [0-0] | 2.4, 2 [0-3] |
| Asthma (100%) | IBS (26%), Depression (20%) | 0.2 | 0.6 | 11.3, 9 [4-16] | 0.4, 0 [0-0] | 1.9, 1 [0-3] |
| IBS (100%) | Depression (29%), Hearing loss (21%) | 0.2 | 0.4 | 10.6, 8 [3-15] | 0.4, 0 [0-0] | 1.2, 0 [0-2] |
| Non-multimorbid |  | 0.1 | 0.2 | 3.7, 1 [0-5] | 0.2, 0 [0-0] | 0.2, 0 [0-0] |
| **Age 45-64 years** |  |  |  |  |  |  |
| Alcohol (62%) | PSM (42%), Pain (28%) | 4.5 | 12.5 | 10.7, 8 [2-16] | 0.5, 0 [0-1] | 2.4, 1 [0-4] |
| Depression (93%) | Pain (53%), Anxiety (31%) | 2.4 | 5.8 | 16.7, 14 [7-23] | 0.6, 0 [0-1] | 5.1, 4 [2-7] |
| Hypertension (76%) | Diabetes (37%), Pain (25%) | 1.6 | 4.4 | 11.5, 9 [4-16] | 0.5, 0 [0-0] | 4.1, 4 [1-6] |
| IBS (40%) | Hearing loss (29%), Pain (28%) | 1.3 | 3.0 | 10.5, 8 [3-15] | 0.5, 0 [0-0] | 2.0, 1 [0-3] |
| Asthma (100%) | Pain (24%), COPD (16%) | 1.0 | 2.7 | 12.6, 10 [5-17] | 0.4, 0 [0-0] | 3.4, 3 [1-5] |
| Non-multimorbid |  | 0.4 | 1.3 | 4.4, 2 [0-6] | 0.2, 0 [0-0] | 0.5, 0 [0-0] |
| **Age 65-84 years** |  |  |  |  |  |  |
| Pain (81%) | CHD (53%), Depression (45%) | 16.2 | 39.2 | 26.3, 23 [14-35] | 1.6, 1 [0-2] | 10.7, 11 [8-14] |
| CHD (54%) | Diabetes (32%), Atrial fibrillation (29%) | 11.3 | 28.8 | 16.8, 14 [6-24] | 1.1, 0 [0-1] | 5.9, 6 [3-8] |
| COPD (57%) | Asthma (49%), Pain (33%) | 9.2 | 25.5 | 15.7, 13 [6-22] | 0.8, 0 [0-1] | 5.5, 5 [2-8] |
| Depression (56%) | Pain (56%), Anxiety (23%) | 8.4 | 20.9 | 17.3, 14 [8-23] | 0.8, 0 [0-1] | 5.9, 6 [3-8] |
| Hypertension (100%) | Diabetes (31%), Pain (27%) | 4.7 | 13.2 | 12.4, 10 [4-17] | 0.6, 0 [0-1] | 4.5, 4 [2-7] |
| Hearing loss (40%) | Prostate disorder (21%), IBS (3%) | 4.4 | 11.1 | 13.2, 11 [5-19] | 0.7, 0 [0-1] | 3.3, 3 [1-5] |
| Non-multimorbid |  | 1.9 | 6.6 | 6.4, 4 [0-9] | 0.3, 0 [0-0] | 1.2, 0 [0-2] |
| **Age 85+ years** |  |  |  |  |  |  |
| CHD (61%) | Atrial fibrillation (53%), Heart failure (49%) | 37.7 | 70.8 | 21.9, 18 [9-31] | 1.5, 1 [0-2] | 8.0, 8 [5-11] |
| Pain (64%) | Depression (41%), Constipation (24%) | 31.1 | 62.9 | 17.3, 15 [7-24] | 0.8, 0 [0-1] | 6.7, 7 [4-10] |
| Asthma (48%) | COPD (48%), Pain (44%) | 28.0 | 56.5 | 19.6, 17 [9-27] | 1.1, 0 [0-2] | 6.9, 7 [4-10] |
| Hypertension (72%) | Hearing loss (39%), Diabetes (18%) | 20.9 | 49.5 | 13.0, 10 [2-19] | 0.8, 0 [0-1] | 4.1, 4 [0-6] |
| Non-multimorbid |  | 13.4 | 36.0 | 6.7, 3 [0-10] | 0.4, 0 [0-0] | 1.3, 0 [0-2] |

Notes: IBS=Irritable bowel syndrome, PSM= Psychoactive substance misuse (not alcohol), CHD=Coronary heart disease, COPD = Chronic Obstructive Pulmonary Disease.

### Multimorbidity clusters and outcomes

For ease of reference we refer to each cluster by its lead or key conditions (i.e. one or three conditions respectively, whose cluster-specific prevalence is highest, and higher than their overall prevalence in their respective age group).

These clusters differ across age strata, both in terms of number of clusters per strata and main components within each cluster (Table 3 and supplementary figures 1-4). The association between multimorbidity clusters and outcomes (service use and mortality) remained significant (p<0·01 in almost all clusters) after stratifying by age strata, controlling for socioeconomic status, BMI and smoking behaviour (supplementary tables 8-11). Results for the distribution of outcomes (e.g. median, interquartile range (IQR)), covariate-adjusted incidence rate ratios for service utilisation and odds ratios (OR) for mortality derived from generalised linear models are available in supplementary tables 8-15 (adjusted covariates were gender, socioeconomic status, smoking status, BMI and age), but with a cautionary note that the relationship between clusters, patient demographics and outcomes should not be interpreted causally.

#### Age strata: 18-44 year olds

Five clusters were uncovered in the 18-44 age strata, whose lead conditions were depression (the most common cluster), pain, asthma, irritable bowel syndrome, and psychoactive substance misuse. Those in the cluster whose three key conditions were depression (100%), anxiety (41%) and pain (31%) were found to have the highest use of primary care consultations (median 12 [5-20] in a year). Those in the cluster whose three key conditions were pain (36%), hearing loss (30%) and hypertension (23%) were found to have the highest hospital admission rates (an average of 0.6 visits in a year) and the highest number of medicines repeatedly prescribed (median1 [0-4] unique drug classes in a year). Highest mortality in this age range was found in the least prevalent (7%) multimorbidity cluster whose three key conditions were psychoactive substance misuse (75%), alcohol problems (42%) and depression (24%) (3.9% mortality in five years). This level of mortality was 18 times higher than that of individuals in the same age range without multimorbidity (0.2%). This cluster was predominantly male (72%), socio-economically deprived (63% in greater deprivation) and with high smoking rates (76% current smokers).

#### Age strata: 45-64 year olds

In the 45-64 age strata, LCA revealed five clusters, whose lead conditions were hypertension (the most common cluster), irritable bowel syndrome, depression, asthma and alcohol problems. Those in the cluster whose three key conditions were depression (93%), pain (53%) and anxiety (31%) had the highest number of primary care consultations (median 14 [7-23] in a year), admission rates (an average of 0.6 visits in a year) and prescribing rates (median 4 [2-7]). As in the younger age strata, the least prevalent multimorbidity cluster (4%) had the highest death rate (13% in five years), its key conditions were alcohol problems (62%), psychoactive substance misuse (42%) and pain (28%). Again, this cluster was characterised by being typically male smokers from areas of high socio-economic deprivation. Pain as a co-morbidity was represented in all the clusters in this age group.

#### Age strata: 65-84 year olds

Six clusters were found in the 65-84 age strata, whose lead conditions were hypertension (the most common cluster), hearing loss, depression, coronary heart disease, chronic obstructive pulmonary disease and pain. The least prevalent multimorbidity cluster (5%), whose key conditions were pain (81%), coronary heart disease (53%) and depression (45%) had the highest use of primary care consultations (median 23 [14-35] in a year), admission rates (an average of 1.6 visits in a year), prescribing rates (median 11 [8-14]) and death rates (39% mortality in five years). Again, this cluster consists of patients with the highest level of deprivation (43%).

#### Age strata: above 85 year olds

The 85+ age stratum was composed of four clusters, whose lead conditions were hypertension (the most common), pain, heart failure and asthma. The majority of patients (58%) fitted within a cluster whose key conditions were hypertension (72%), hearing loss (39%) and diabetes (18%). The cluster with the majority of patients had the lowest mortality (50% five-year mortality), as well as the least number of conditions (median 3 [2-4] morbidities), and the least health care utilisation (roughly half the GP contacts, hospitalisations and medicines on repeat prescription of the cluster whose lead condition was “coronary heart disease”). The cluster with the highest mortality, GP contact, hospitalisations and repeat prescriptions comprised a trio of cardiac conditions: coronary heart disease, atrial fibrillation and heart failure.

### Validation of cluster morbidity profiles

As well as validating the clusters by their association with patient characteristics and outcomes, the similarity of multimorbidity clusters were compared between the training set (80% of patients, N=90,571) and the test set (the other 20% of patients, N=22,640). Results are summarised below, and given in full in supplementary material section 4. Measures of cluster quality (i.e. entropy) were found to be consistent between the training and test sets.

As the training set contained more disease patterns, the derived clusters were more comprehensive. The test set (with fewer patients) contained fewer disease patterns and therefore we expected the derived clusters to be a subset of those in the training set. Indeed, validation of cluster profiles showed that every cluster in the test set found a match in the training set. Some clusters were particularly robust (had the smallest JSD and the highest Pearson’s correlation coefficient), for instance, those in the largest age strata (65-84 age strata, N = 49,494), and clusters whose lead condition was depression, psychoactive drug misuse or alcohol problems. Clusters with a less clear match were the cluster in the 18-44 age strata whose lead condition was asthma.

## Discussion

### Summary of results and comparison with other studies

This study identified and validated clusters of multimorbid patients using a novel patient-centred approach. In summary, we identified twenty patient clusters across four age strata. In the younger age-strata (18-44; 45-64), the clusters with the highest mortality (18 times higher than the non-multimorbid group in 18-44 year olds) comprised psychoactive substance abuse in combination with alcohol problems. The clusters with the most contact with general practice in people aged under 65 comprised depressions, anxiety and pain. In 65-84 year olds, the cluster with the highest mortality and highest health service use (GP contact; hospitalisations; repeat prescriptions) comprised pain, coronary heart disease and depression, and in people aged 85 or over, it comprised heart failure, coronary heart disease and atrial fibrillation. The most common cluster in 18-44 year olds was centred around depression, but in all other age groups, they were centred around hypertension. In the oldest age group, this hypertension-centred cluster was associated with the best survival and lowest health service use among multimorbid patients. Pain featured in thirteen of the clusters.

In this study, unlike most previous analyses of multimorbidity, we have defined clusters in terms of patients rather than diseases^9,10^. As a result, we have identified novel clusters that have practical implications for service delivery. The high mortality of the cluster of psychoactive substance and alcohol misuse warrants attention, and the descriptive epidemiology of this cluster (male, smoker, under age 65, high socio-economic deprivation, relatively low service use) gives pointers as how the poor health might be addressed. Conversely, we found that the commonest cluster in people aged 85 and over (58% of patients with multi-morbidity in this age group) was associated with the least health service use and the lowest mortality. This gives a more nuanced perspective on the association of multimorbidity with age that has been widely reported^7^, in that it suggests that in the oldest age group, multimorbidity *per se* may be less important, although numerically most common. Our age-stratified approach also enables different patterns of co-morbidity to be identified. Thus, in younger age groups, clusters focused around mental health are associated with most GP contact, in people aged 65-84, a cluster of mental health and coronary heart disease is associated with most GP contact, and other indicators of health service use, whereas in people aged over 85, the cluster representing most health service contact is dominated by cardiovascular conditions. In terms of relative importance of single conditions within multimorbid clusters, the predominance of mental health conditions and hypertension has been identified in previous work^9,10,17,26^. A novel finding in our work is the inclusion of pain in many of the clusters we identified. There are several possible explanations for this, including the conditions studied, as well as the different methodological approach we took.

### Strengths and limitations

The robust identification of such clusters would not have been possible without the novel use of age-stratification, patient-level clustering (not requiring all patients to have identical lists of conditions) and validation with held-out data. This is the largest scale application of age-stratified latent class analysis to multimorbidity, both in patient numbers (above 100,000) and the number of conditions (38)^10^. By including younger patients and stratifying by age we see how multimorbidity clusters differ over the lifecourse. Combining this with the release of reproducible analysis scripts is an approach which we recommend for future multimorbidity clustering efforts. Our systematic approach including all 38 conditions from Barnett et al., ^7^ age-stratification, clustering and outcomes, was necessary to handle the complexity of multimorbidity in healthcare.

This study suffers from typical limitations of electronic health record research in that they rely on routine coding within the healthcare system including residual confounding and variable CPRD data quality. Wherever practically feasible we have taken steps to address these, e.g. the careful design of codelists, relying on variables with low missingness and adjusting for key variables. While the clustering approach used (LCA) is a robust probabilistic approach, results may differ subtly if other approaches are used. Validation of latent clusters also requires further research where a larger sample size for the test set, perhaps from another database or country, can strengthen the validation. Despite this, given the large and representative sample, the consistency of results both internally, across age strata and with existing literature, we are confident in our main results. Finally, multimorbidity evolves over time, but we only use longitudinal data to extract conditions in 2012 and service use and mortality outcomes.

### Conclusion and policy implications

These multimorbidity clusters highlight major targets for public health and healthcare, giving a more nuanced understanding of multimorbidity than the work of Barnett et al^7^ and others which rely more on simple disease counts. The 18-fold higher mortality of younger multimorbid patients with psychoactive substance misuse is a clear case of an unmet need. Improving outcomes for this neglected patient group is likely to be feasible given that their risk factors (drug use, smoking, deprivation) are potentially amenable to intervention. Conversely, the fact that the majority of older multimorbid patients have relatively low service use and mortality has implications for the design of health services. More generally, the fact that chronic pain is a key feature of many multimorbidity clusters suggest that it is important to manage pain within the context of multimorbidity rather than in its own right. Similarly, our findings add to the evidence showing the importance of mental health in multimorbid patients, justifying the push for parity of physical and mental health within the healthcare system.

While patients with multimorbidity account for an ever increasing proportion of healthcare need and provision^1,4,5,7^, no existing interventions have shown convincing evidence of benefit in improving important outcomes^8,27^. Our findings fit with the suggestion from Salisbury et al.,^8^ that one reason for the failure of previous interventions is that multimorbidity is heterogeneous, with very different diseases, needs and outcomes in different groups of patients. Our findings support the proposal that interventions to improve outcomes in multimorbidity may be more appropriately targeted on distinct types, and we have systematically highlighted groups of patients where tailored approaches could be attempted.

## Data Availability

CPRD-GOLD is available to approved researchers for projects approved by the CPRD Independent Scientific Advisory Committee

## Supplementary files

Supplemental file 1 – ISAC protocol (16_057RA2)

Supplemental file 2 – Supplementary.docx

Supplemental file 3 – STROBE checklist

## Acknowledgements

We acknowledge CPRD @ Cambridge for developing and sharing disease definitions, and Dr Jennifer Quint (Imperial College London) for permission to use and share a codelist for smoking status. We also acknowledge the valuable statistical discussions with Dr. Robert Goudie and Dr. Paul Kirk at MRC Biostatistics Unit, University of Cambridge. This study is based in part on data from the Clinical Practice Research Datalink obtained under licence from the UK Medicines and Healthcare Products Regulatory Agency. The data is provided by patients and collected by the NHS as part of their care and support. ONS is the provider of ONS mortality data used in this study. ONS and HES data copyright © (2018), re-used with the permission of The Health & Social Care Information Centre. All rights reserved. The interpretation and conclusions contained in this study are those of the author/s alone.

